# A Translational Neuroscience & Computational Evaluation of a D1R Partial Agonist for Schizophrenia (TRANSCENDS): Rationale and Study Design of a Brain-Based Clinical Trial

**DOI:** 10.1101/2025.04.18.25326082

**Authors:** Clara Fonteneau, Zailyn Tamayo, Ally Price, Lining Pan, Yvette Afriyie-Agyemang, Shriya Agrawal, Audrey Butler, Courtney Cail, Monica Calkins, Srinivas Chakilam, Kimberlee Forselius-Bielen, Geena Fram, Allea Frazier, Roberto Gil, Preetika Govil, David L. Gray, Jack Grinband, Ruben C. Gur, Natalka K. Haubold, Zachary Heffernan, Larry Kegeles, Christian Kohler, Chenyang Lin, Jing Lu, Megan Mayer, Phuong Pham, Greg Perlman, Masih Rahmati, Mohini Ranganathan, Nicole P. Santamauro, Christopher T. Schutte, Alexandria Selloni, Jared Van Snellenberg, Toral Surti, Daniel H. Wolf, Catrin Zharyy, TRANSCENDS Group, Anissa Abi-Dargham, Raquel E. Gur, Jeffrey A. Lieberman, Joshua T. Kantrowitz, Alan Anticevic, Youngsun T. Cho, John H. Krystal

**Author notes:** Contributed equally to this work (Clara Fonteneau, Zailyn Tamayo). Contributed equally to this work (Alan Anticevic, Youngsun T. Cho, John H. Krystal).

## Abstract

Despite decades of research, cognitive impairment remains a critical untreated symptom for many patients with schizophrenia. One way to accelerate the development of pro-cognitive therapies for schizophrenia is to evaluate compounds using biomarker approaches tailored to relevant neural mechanisms. While D1/D5 receptor (D1R/D5R) agonism has been extensively studied in neuroscience, its therapeutic potential for cognitive impairment in schizophrenia remains untapped. The Translational Neuroscience & Computational Evaluation of a D1R Partial Agonist for Schizophrenia (TRANSCENDS) clinical trial tests this mechanism using a ‘target engagement’ approach. Multiple, double-blind doses of a D1/D5R partial agonist were administered in advance of a functional neuroimaging (fMRI) session that deployed a cognitive paradigm explicitly designed to capture a translational micro-circuit mechanism underlying spatial working memory in patients with schizophrenia. Specifically, this study will assess whether the D1R/D5R partial agonist CVL-562 induces a dose-dependent engagement of spatial working memory circuits in schizophrenia using fMRI. This design, and the use of spatial working memory neural circuits as a dependent measure, was selected on the basis of a translational and computational understanding of prefrontal micro-circuitry and a mechanistic understanding of the role of D1R/D5Rs in schizophrenia. To enhance data integration and scalability, TRANSCENDS employs an automated informatics framework for seamless neuroimaging data sharing and electronic clinical data capture. This ensures high-standards for regulatory compliance, data quality, and data sharing across sites, improving aspects of current clinical trial data management. We share the study design and approach with the goal of advancing future pro-cognitive drug development and strategies for developing mechanistically-driven biomarkers in psychiatry.

## Introduction

The NIMH-funded multi-site clinical trial **TRA**nslational **N**euro**S**cience & **C**omputational **E**valuatio**N** of a **D**1R Partial Agonist for **S**chizophrenia (TRANSCENDS) addresses the NIMH Program Announcement for National Cooperative Drug Discovery/Development Groups (NCDDG). This study aims to advance the translation of promising compounds by establishing proof-of-concept trials to demonstrate engagement of targeted neural circuits (i.e., target engagement). TRANSCENDS seeks to identify neuroimaging biomarkers of D1R/D5R engagement to accelerate development of D1R/D5R agonists for treating cognitive impairments in schizophrenia, which are linked to greater functional disability and are a key public health concern (1, 2).

All D1R agonists studied to date activate both D1 and D5 receptors. D1R/D5R agonism is one of the most intensively studied therapeutic mechanisms from a basic neuroscience perspective, but least understood mechanisms from a clinical perspective. Patricia Goldman-Rakic first identified procognitive effects of D1R/D5R agonism in non-human primates (3). Yet, D1R/D5R treatments for schizophrenia have not been successfully developed for reasons that may include: i) only very recent development of D1R/D5R agonists with good CNS bioavailability; ii) steep inverted-U dose-related effects of D1R/D5R agonists on working memory; which makes optimal dose selection difficult (4–7); iii) acute behavioral effects of D1R agonists that may not be indicative of long-term effects in D1R-sensitized networks (8, 9); iv) chronic antipsychotic treatment downregulating D1Rs and complicating the optimal dosing of D1R agonists in patients (6); v) specific predictions about D1R/D5R agonist effects from preclinical and computational studies (10–12) that have not borne out in clinical studies using neuropsychological tests that may not be optimized to detect D1R agonist effects (13, 14), and, finally; vi) illness phase possibly influencing D1R/D5R agonist response (15, 16). Collectively, these reasons suggest that the pro-cognitive potential of a D1R/D5R agonist in humans has yet to be optimally tested.

The translational framework underlying this study posits the hypothesis that D1R/D5R agonists improve cognition, specifically, working memory (WM), by restoring deficient inhibitory tuning of executive cortical networks in patients with schizophrenia (Figure 1A). In patients with schizophrenia, Abi-Dargham (TRANSCENDS site PI) and colleagues have previously shown deficits in PFC dopamine release and upregulation of D1R/D5Rs in association with WM impairments, implicating D1R/D5R signaling impairments in cognitive dysfunction in schizophrenia (17–19). In non-human primates, Goldman-Rakic, Arnsten, and colleagues showed that deficits in D1R/D5R signaling reduce neural tuning during WM (20, 21). In turn, D1R/D5R agonists restored neural activity associated with spatial WM (sWM) and suppressed the activity of spatially-specific pyramidal neurons to non-preferred spatial locations (i.e., reduced the noisiness of spatial tuning) (5, 22, 23). Using a delayed response sWM task, a number of studies have showed that D1R/D5R agonists attenuate the sWM disruption produced by haloperidol administration (24), amphetamine sensitization (8, 25, 26), ketamine administration (27, 28), and aging (5, 8). Computational models of this circuit show that, while enhancing tuning, D1R/D5R agonism stabilizes recurrent neural activity and reduces effects of distractors (29). Further interactions between D1R and glutamatergic signaling, especially through NMDA receptor antagonism, as modeled by ketamine, are likely critical for supporting cortical microcircuit mechanisms of WM (30–39).

**Fig. 1.**
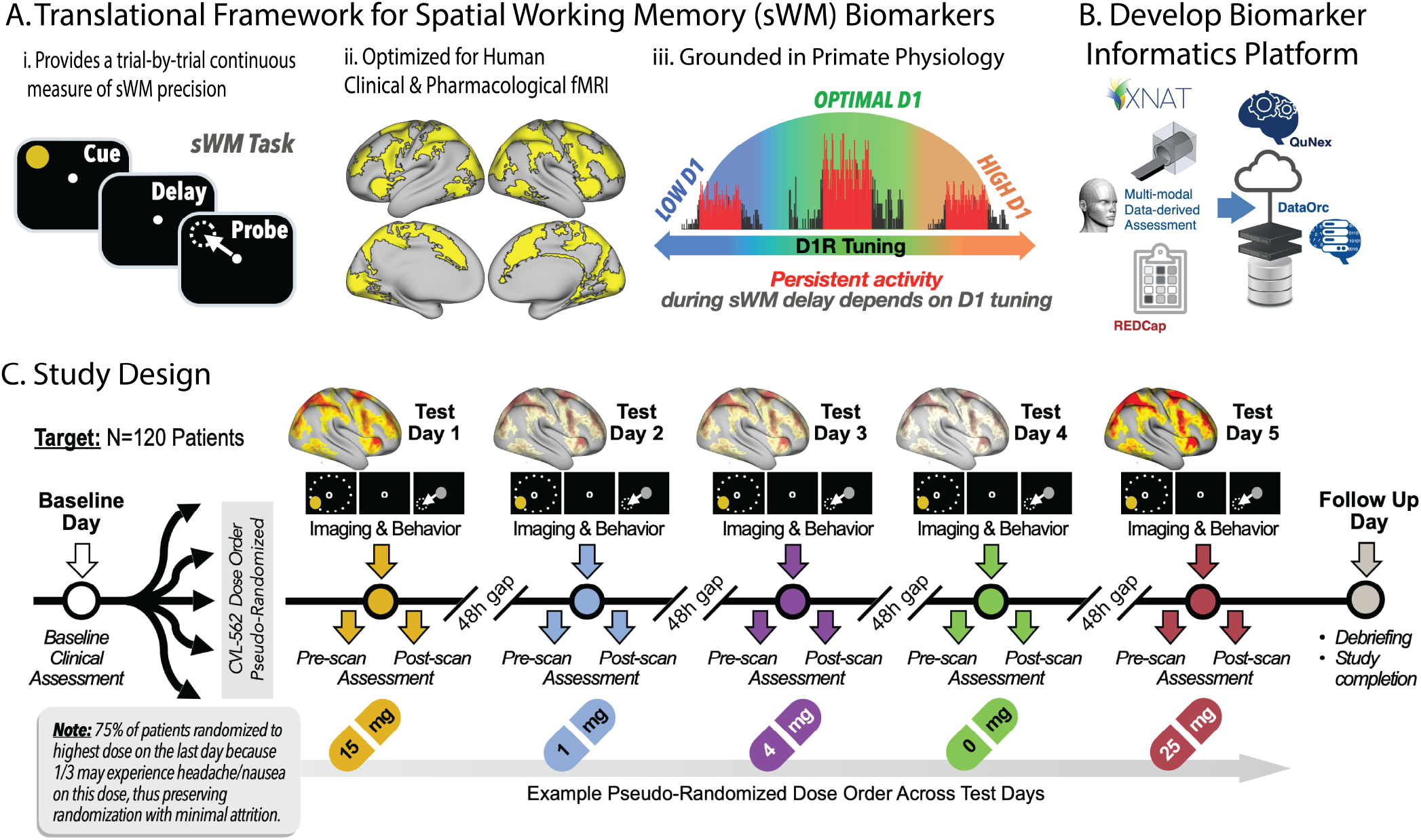
TRANSCENDS Study Overview. A. Translational Framework for Spatial Working Memory (sWM) Neural Biomarkers Overview. The translational framework designed for the TRANSCENDS study is that D1R/D5R agonists improve working memory in schizophrenia by restoring deficient inhibitory tuning within cortical microcircuits. A) i) Spatial working memory task from Cho et al. (43). The task has 4 conditions: Motor, Spatial Working Memory (No Distractor), Distractor Near (20° or 40° distance), Distractor Far (60° or 80° distance), and provides a continuous measure of sWM precision; ii) the sWM task is optimized for probing human cognitive circuits using fMRI. Panel refers to Figure 2A from Cho et al (44) showing regions activated by sWM across encoding and delay epochs, including frontoparietal circuitry; iii) sWM task is grounded in studies of primate physiology, which suggest that optimal stimulation of the D1R occurs in a dose-dependent, inverted-U manner to support sWM (adapted from Goldman-Rakic, Arnsten, and colleagues (5, 22, 23)). B. The TRANSCENDS informatics infrastructure is designed to provide automated data quality assurance, interoperable electronic data capture (EDC), validated systems integration, seamless data sharing, and an analytics platform for biomarker readout. C. Overview of the clinical trial design, which uses a double-blind multi-dose strategy across five separate test days to examine dose-dependent effects of D1R/D5R partial agonism on working memory neural circuits.

Building upon the evidence that D1R mechanisms are crucial for supporting working memory, TRANSCENDS aims to test whether CVL-562 (previously PF-06412562), a dopamine D1/D5 receptor partial agonist, affects working memory neural circuits in schizophrenia patients using a spatial working memory (sWM) task. The primary aim is to assess dose-related neural circuit targets of this compound, while secondary aims will quantify drug effects on sWM precision and functional connectivity.

Clinical trials that leverage neuroimaging biomarkers are paramount for identifying promising treatments, yet the current lack of field-wide standards in informatics and data science harmonization can limit the deployment of these types of studies, which rely heavily on collecting and organizing large amounts of data. In particular, this is a key challenge for the integration of multi-site, large-scale neuroimaging data collected as part of clinical pharmacological studies. To help improve this aspect of neuroimaging biomarker development, we developed a data orchestration framework (DataOrc) that combines the rigorous regulatory requirements of clinical trials, with the technical complexities needed for rapid and efficient neuroimaging transfer and storage (Figure 1B). This pharmacologic neuroimaging infrastructure is designed to provide automated data quality assurance, interoperable electronic data capture (EDC), validated systems integration, seamless data sharing, and an analytics platform for biomarker readout. Specifically, this architecture features the integration of three ecosystems to enable full EDC-driven clinical trial deployment: i) XNAT, an extensible open-source imaging informatics software platform (40); ii) A CFR Part 11 compliant REDCap database (41); and iii) the Quantitative Neuroimaging Environment & ToolboX suite that enables cloud-based containerized neuroimaging biomarker analytics (42).

This brain-based clinical trial design, coupled with our informatics and data orchestration framework, provides an example for future studies of neuroimaging biomarkers of pharmacological treatment. Such an ecosystem considers the importance of supporting rational approaches to treatment development, as opposed to serendipitous drug discovery, for example, in pursuit of identifying biomarkers that are clinically actionable. This paper describes the rationale and design of the TRANSCENDS study with the aim of highlighting its unique scientific and technological aspects.

### Study Design

TRANSCENDS is a Phase II brain-based clinical trial developed to test whether CVL-562, a D1R/D5R partial agonist novel compound, affects working memory neural circuits in patients with early episode schizophrenia (Figure 1C). Patients were recruited from four centers experienced in recruiting and enrolling patients with early-episode psychosis into research (Columbia University/Research Foundation for Mental Hygiene (RFMH), SUNY Stony Brook (SBU), University of Pennsylvania, Yale University) over a 2-3 year period. Eligible patients participated in research activities across 7 study visits within a approximately 1-2 months period (Figure 1C). Five of these study visits involved the administration of CVL-562 or placebo. Each test visit was separated by at least 48 hours (six half-lives of CVL-562). See Table 1 for the schedule of all events during each visit.

**Table 1.**
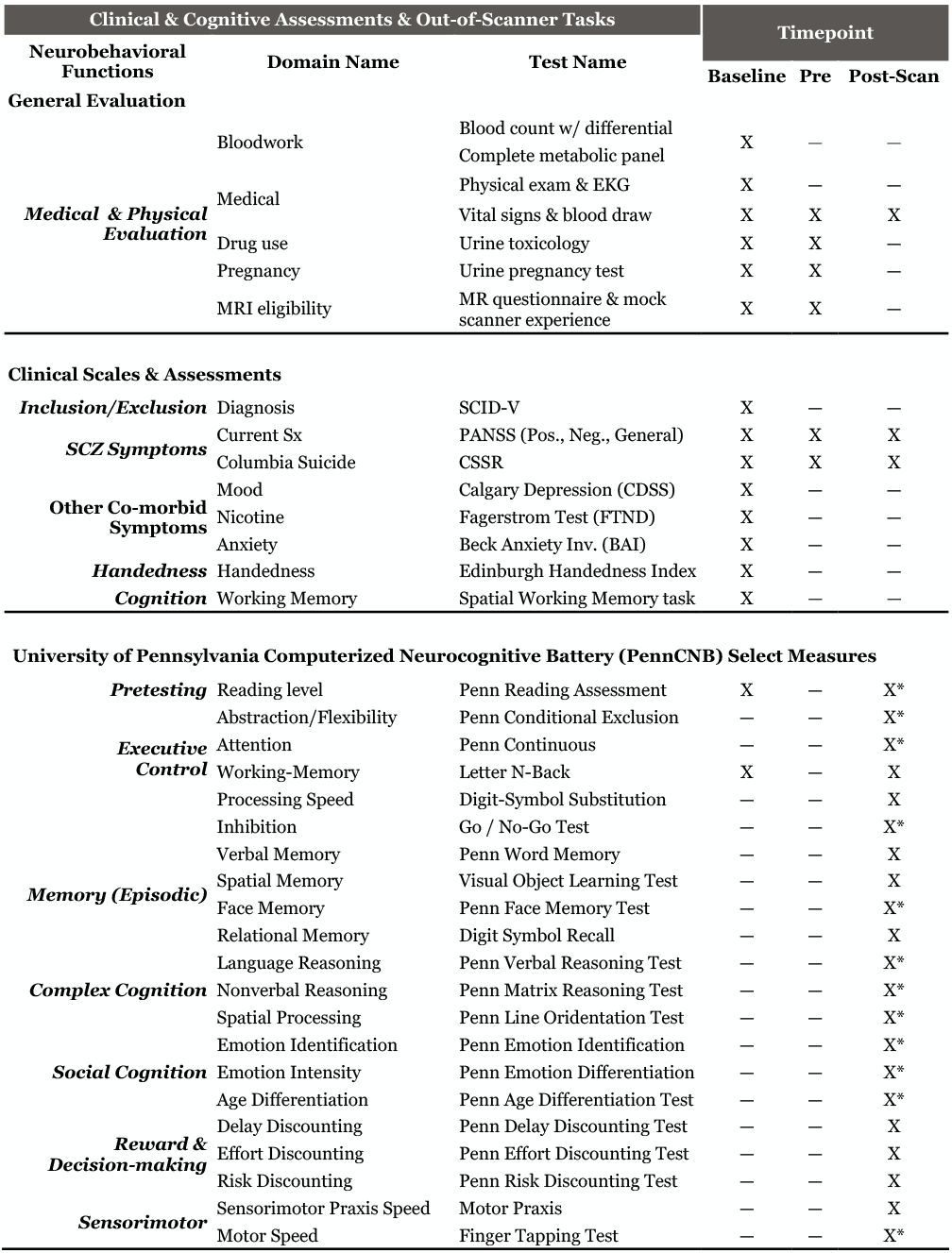
Schedule of Events [SoE]. Abbreviations: Structured Clinical Interview for DSM-V (SCID-V); Positive and Negative Syndrome Scale (PANSS); Columbia Suicide Severity Rating (C-SSR); The Calgary Depression Scale for Schizophrenia (CDSS); Fagerstrom Test for Nicotine Dependence (FTND); Beck Anxiety Inventory (BAI). * represents assessments done at Visit 2 and 6 only.

After a brief phone screen that included a description of the study, patients were invited to participate in an in-person baseline assessment visit that determined eligibility in the study. Assessments included a semi-structured clinical interview (SCID-5, Schedule Clinical Interview for DSM-5), health history, concomittant medications, urine toxicology, bloodwork, EKG, and cognitive testing. Cognitive testing included the Penn Reading Assessment (PRA), a trainer task designed to teach participants to centrally fixate during the spatial working memory task, and a practice version of the spatial working memory (sWM) task. The latter two tasks were included to ensure that participants understood, and were adequately able to follow, instructions for the sWM task (Figure 2) (34, 43–47).

**Fig. 2.**
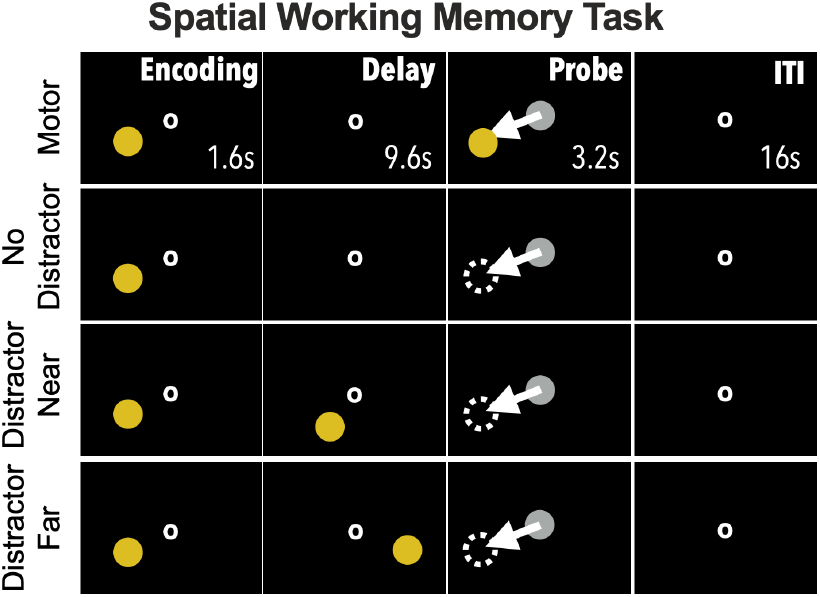
Spatial Working Memory Task. The task has 4 conditions (20 trials each): Motor, Working Memory (No Distractor), Distractor Near (20° or 40°), Distractor Far (60° or 80°). In each condition, a yellow circle cues the spatial location to be encoded. In the working memory and distractor conditions, participants have been instructed to keep in mind the spatial location (delay epoch), which they indicate using a joystick to move the grey circle to the remembered location (probe epoch). In the control motor condition, no working memory is required, and the cued location re-appears for the participant to place the grey circle as close as possible to it. In both distractor conditions, the distractors appear half-way through the delay epoch. The outcome measure is the angular distance between the cued location and probe placement. This task provides a trial-by-trial continuous measure of precision.

Upon reviewing all of the information collected at the baseline assessment visit, the study physician at each site signed off on an eligibility checklist in REDCap to document eligibility for the additional activities of the study. Once deemed eligible, participants participated in 5 test visits that involved a double-blind dose of CVL-562 or placebo, followed by a functional magnetic resonance imaging (fMRI) scan. Prior to drug administration at each test visit, vitals signs, a pre-dose Positive and Negative Symptoms Scale (PANSS) interview, Columbia-Suicide Severity Rating Scale (CSSRS), urine toxicology and concomitant medications were assessed. A lifestyle questionnaire was administered to understand basic events that occurred in the hours before the scan and a brief refresher session on the sWM task was presented to participants to remind them of the task instructions. After these procedures, the test visit medication was administered, and the subject entered the MRI scanner. Scanning sequences included T1-weighted and T2-weighted structural MRIs, four functional BOLD runs of the sWM task, one resting-state BOLD run, one pseudo Arterial Spin Labeling (pASL) sequence, and one functional BOLD run of a visual flashing checkboard task. One diffusion-weight image (DWI) was additionally collected at one of the study visits. Scanning was timed such that functional imaging of the sWM task coincided with peak CVL-562 plasma levels (approximately 75 minutes following medication administration). Serum levels of CVL-562 were collected prior to and following imaging. Vital signs, PANSS, CSSRS, and PennCNB were done immediately following exit from the scanner. All participants were observed for adverse events for at least 2 hours following dosing and were provided transportation home in order to avoid driving. After completion of the 5 treatment visits, participants were scheduled for a final in-person follow-up visit that involved bloodwork and an exit interview with debriefing of the study.

### Rationale for Schedule of Events

#### Selection of Drug

Prior D1R/D5R agonists studied in schizophrenia included SKF38393, a weak partial agonist with poor CNS bioavailability that produced ambiguous results, and dihydrexidine/DAR-100A, a full agonist at both receptors, with very steep inverted U dose-related effects and unclear benefits (14). Full agonists of the D1R have been shown to be limited by tachyphylaxis and tolerance (48). In contrast, CVL-562 is a partial agonist with high selectivity for D1R/D5R, oral bioavailability and moderate binding affinity for both recombinant human D1R and human D5R (49–51). The partial agonist activity of CVL-562 at D1R may protect against detrimental cognitive effects related to overstimulation of D1R. Relatedly, direct impairment of cognition was not observed in the Pfizer Phase IB SCZ trial of CVL-562 (13). Unfortunately, the initial Phase IB study of CVL-562 in schizophrenia was negative and did not provide a clear guidance for dose-selection (13). Non-human primate studies conducted by Pfizer describe dose-related improvements in sWM (28), and with chronic intermittent administration, even lower doses of D1R/D5R agonists that were without initial benefit also exhibited procognitive effects (30). Notably, Phase I studies have shown CVL-562 to be safe and well-tolerated in humans (52–55). Our study design incorporates a wide range of doses (0, 1, 4, 15 and 25mg) in order to identify neural responses to specific doses and allow us to map individual pharmacologic dose-response profiles. The particular doses have been chosen based on prior work using this drug, and in order for the group to have a range of doses to chart a purported inverted-U curve of drug dosing with respect to impact on cognitive functioning (7). The 1 mg dose was included in order to identify those participants who are the most sensitive to D1 partial agonism. As we intend to chart the full dose-response for each participant, the selected doses will allow us to catch those participants who are either more or less sensitive to D1 partial agonism, as well as chart differential pharmacodynamics curves.

### Selection of Behavioral and Cognitive Tasks

#### Spatial Working Memory Task

This study builds on prior literature describing the mechanisms through which D1R/D5R agonists enhance spatial working memory (sWM): 1) they stabilize persisting neural representations over temporal delays and in the face of distraction, 2) they optimize the inhibitory tuning of neural representations, sculpting more precise sWM representations (7, 9, 56). To optimize both features, we use a task that is a direct translation of the spatial delayed response task developed by Funahashi and Goldman-Rakic and employed in subsequent studies of D1R/D5R agonists in the Goldman-Rakic laboratory and others (3, 7, 45, 57). The human version of the task involves having people move their eyes (58, 59) or direct a joystick to the precise spot where a target stimulus was presented (43, 44), both in the presence of, and absence of, distractors of varying proximity to the target (34, 43, 44, 58). In this task, patients with schizophrenia show relatively greater impairment in spatial working memory precision, with additional impairments seen in response to increasing delay epochs and the presence of distractors (34, 43, 60). This sWM framework builds on rich evidence from non-human primates implicating specific neurophysiologic mechanisms supporting sWM (61–63), which can also be leveraged for understanding clinical deficits via computational modeling (33, 64, 65). See Figure 2 for details on the sWM task used.

#### PennCNB

Out-of-scanner neurocognitive functioning is assessed using select measures from the University of Pennsylvania Computerized Neurocognitive Battery (PennCNB) (https://penncnp.med.upenn.edu), which was developed and optimized by consortium co-investigator Dr. Ruben Gur. Cognitive domains in the PennCNB include: Abstraction and Flexibility; Attention; WM; Episodic Memory; Language Reasoning; Spatial Processing; Sensorimotor Processing Speed; Motor Speed; Emotion Identification, providing additional measures to test specificity/generalizability of in-scanner WM results. Alternate versions are used longitudinally to control for practice effects, which is also explicitly controlled for, given the longitudinal design (66).

#### Selection of Clinical Markers

To study clinical predictors and moderators of CVL-562 effects on neuroimaging biomarkers, we extensively characterized patients at the baseline assessment, and then conducted focused clinical and neuropsychological assessments, including the Positive and Negative Syndrome Scale (PANSS), Columbia Suicide Severity Rating Scale (C-SSRS), and Lifestyle questionnaire on each test day.

#### Selection of Imaging Markers

We hypothesize that the D1R/D5R partial agonist CVL-562 has pro-cognitive effects in schizophrenia by restoring inhibitory tuning of prefrontal cortical activity, thereby increasing sWM behavioral precision and reducing the impact of distractors (23, 67). The overarching goal of this trial is to test whether D1 partial agonism is a viable pro-cognitive mechanism in human participants with schizophrenia. To this end, our primary endpoint will be neuroimaging, and we will test whether D1 partial agonism affects sWM circuits, with improved sWM performance and the identification of a subset of those who respond to CVL-562 as secondary endpoints. Using these as secondary endpoints could facilitate subsequent full-scale clinical trials if our study demonstrates single-dose effects of CVL-562 on associated neural circuits.

#### Selection of Genetic and other Blood Based Markers

Collecting blood work for genetics was optional on the part of the participant (i.e., opt-in). The use of genetic materials will be exploratory and used in relation to imaging and treatment findings. All blood collected for genetics analysis was shipped to and stored at Yale University. Any extracted genetic data will be made available through the National Institutes of Health database of Genotypes and Phenotypes (daGaP).

Serum levels of CVL-562 were obtained prior to and following imaging. Following collection, blood was allowed to clot, and then centrifuged to separate the components. The serum was carefully aliquoted for freezing (-80F) and analysis. Serum level measurements were done at Columbia Unviersity’s Irving Medical Center, following validation protocols provided by Cerevel.

### Study Aims

This clinical trial will focus on sWM-related functional neuroimaging as the primary outcome measure, in order to evaluate whether CVL-562 exhibits a dose-related neural effect in patients with schizophrenia (<10 years of psychosis duration). Predetermined secondary endpoints will focus on the drug-related effects on: 1) The proportion of participants with a BOLD signal response during the sWM task; 2) Performance during the spatial working memory task; 3) Associations between BOLD signal and spatial working memory performance; 4) Functional connectivity of the frontoparietal network with the rest of the brain during the sWM task; 5) Resting state global brain connectivity; 6) Spatial similarity of resting state global brain connectivity with transcriptomic maps. A final, exploratory endpoint will test the association of genetic variants with BOLD signal changes. Collectively, this translational biomarker study informs a high priority experimental treatment mechanism identified by the NIMH MATRICS Initiative by testing the engagement of cognitive circuits by D1R partial agonism in humans with schizophrenia (68).

### Study Participants

#### Overall procedure across sites

This trial is conducted according to the FDA guidelines and approved by Institutional Review Board at each clinical site. Recruitment leveraged an established pipeline across study sites, primarily drawing patients with schizophrenia-spectrum disorders from specialized early psychosis programs. Additional participants were referred from clinics, inpatient units, ERs, and intensive outpatient programs, supplemented by advertising and community outreach. The study aimed to enroll 120 adults, targeting 100 completers, with efforts to ensure gender and racial diversity. Eligibility was determined by specific inclusion/exclusion criteria, including a psychosis duration of fewer than 10 years (Table 2).

**Table 2.** Summary of major inclusion and exclusion criteria for participants in the TRANSCENDS study.

| Major Inclusion/Exclusion Criteria |
| --- |
| **Diagnosis:** DSM-5 diagnosis of schizophrenia, schizoaffective, or schizophreniform disorder within 10 y. of illness onset. |
| **Age:** Between 18 and 45 years of age. |
| **Mental Status:** Have capacity to consent. |
| **Cognitive:** Ability to follow task instructions and perform necessary related motor functions and IQ $\geq 80$. |
| **Medical:** No major medical conditions or active treatment. Clinically stable treatment for at least 2 months prior. No intention to become pregnant and agreement to use reliable method of birth control during the study period. |
| **Psychiatric:** No history of ADHD pre-morbid to the onset of psychosis or other psychiatric illnesses that may be accompanied by cognitive impairment. |
| **Neurological:** No significant head injury (severe concussion, hospitalization) or neurological disorder. |
| **Language/Vision:** Able to read and speak English at 8th grade level and no visual disturbance that cannot be corrected during fMRI. |
| **MR Contraindications:** No metal implants, pacemaker or any other MRI contraindication |
| **Substances:** No recent (past 3mo) comorbid moderate/severe substance use disorder besides nicotine or positive urine toxicology testing for any substance other than marijuana or those prescribed for medical reasons. |
| **Current Antipsychotic treatment:** Stable psychotropic medication regimen for at least 3 weeks prior to enrollment and throughout the study. Cannot be treated currently with any of the following: Olanzapine, clozapine, ziprasidone or asenapine, in order to avoid prominent D1 receptor effects. |
| **Drug-Drug Effects:** No other P450 enzyme inhibitors or inducers |

#### Rationale for Selected Patients

This clinical trial employs two precision medicine strategies: assessing patients on WM impairment and limiting recruitment to patients early in their course of illness. 1) WM Impairment: D1R/D5R agonists are targeted for SCZ because in these patients, deficits in dopamine release and compensatory upregulation of D1R/D5Rs are associated with WM impairments (17–19). Thus, to formally test whether baseline WM impairments are associated with differential responses to the study drug, all participants were measured on the PennCNB letter n-back task during the baseline assessment visit. Individual baseline performance on this task can be compared with drug response during analysis. 2) Illness Phase Specificity: D1R/D5R agonists enhance inhibitory tuning of PFC pyramidal neurons (30), reducing neuronal firing to non-preferred stimuli, i.e., suppressing noise (7). We have previously hypothesized that drugs that have this effect may be relatively more efficacious early in the course of schizophrenia (35), when deficits in inhibitory tuning of PFC pyramidal neurons may be at its most prominent. Further, we had suggested that drugs that attenuate functional connectivity would lose their efficacy in chronic illness because these drugs would exacerbate the damaging impact on network function of illness progression-related loss of grey/white matter and synaptic connectivity (69–73). This hypothesis is supported by early course schizophrenia studies that reported resting-state fMRI (rsfMRI) functional hyper-connectivity relative to later illness stages (74, 75). Importantly, focusing on the early-course illness period reduces the impact of progressive synaptic loss due to advancing illness and the possible cumulative and complex impact of antipsychotic treatment. The notion that drugs that enhance inhibitory tuning might work preferentially early in the course of SCZ is supported by findings with an mGluR2 agonist, which ameliorates symptoms at moderate doses in early course patients (3 yr), but which worsens symptoms at high doses in patients with chronic (>10 yr) schizophrenia (76).

#### Concomitant Medications

Participants were allowed to be treated with psychiatric and medical medications as long as dosing was stable for 3 weeks. Antipsychotics allowed in the study were limited to agents with low D1R affinity, thereby excluding olanzapine, clozapine, ziprasidone and asenapine (77, 78). CVL-562, as well as some of the allowable antipsychotics in this study are metabolized by the P450 enzyme, CYP3A4. Therefore, both strong and moderate inducers (eg, carbamazepine) and inhibitors (eg, ketoconazole, valproate) of CYP3A4 were excluded during this study, as well as the 10 days prior to the initial visit.

### Drug Randomization Protocol

Participants were randomized to the order of doses of CVL-562 (1 mg, 4 mg, 15 mg, or 25 mg) or placebo across 5 study visits. Randomization occurred after the site PI confirmed eligibility for full study procedures. The randomization schema assigned 75% of patients to the highest dose on the last visit, with 25% of patients receiving the highest dose on one of the other four visits. Only the highest dose (25 mg) was be subject to this pseudo-randomization strategy; all other doses were randomly distributed. This randomization strategy was intended to minimize the impact on study completion, as the association of the 25 mg dose with nausea in approximately 30% of participants may unblind participants, with subsequent discomfort leading to study discontinuation. The randomization of dose order will also allow for separation of practice effects, as we can test to ensure that drug effects are stronger than practice effects. Randomization was handled by a statistician and known only to them and the pharmacist.

### Consortium organization

#### Team Coordination activities

The Team leadership established eight working groups (WGs), each led by a designated lead and including representatives from all sites (Penn, Columbia/RFMH, SBU, Yale) and the NIMH advisory team. These WGs cover regulatory, study design, neuroinformatics, assessments, biomarker development and publication strategy. WGs meet weekly, reporting progress in Executive Team Meetings, while Steering Committee and Project Management Team meetings occur weekly via Zoom to oversee study operations.

#### Data Safety Monitoring Board (DSMB)

The Data Safety Monitoring Board (DSMB) meets annually to review study progress, with summaries provided three weeks in advance. The DSMB assesses serious adverse events (SAEs) within 7 days, reviews adverse events (AEs) annually, and monitors recruitment feasibility. Recruitment reports are provided every six months, with an optional meeting as needed.

### Automated and Scalable Framework for a Brain-based Clinical Trial

#### Direct electronic data capture

To support this brain-based clinical trial, we developed a flexible, inter-operable cloud-based informatics solution that enables data aggregation, harmonization, processing, analytics for primary outcome readout and rapid NIMH Data Archive (NDA) data sharing. Specifically, this architecture features the integration of four ecosystems to enable full EDC-driven clinical trial deployment: i) XNAT, an extensible open-source imaging informatics software platform (40); ii) A CFR-Part 11 compliant REDCap database housed by the Yale Center for Clinical Investigation (YCCI) (41); iii) QuNex, the Quantitative Neuroimaging Environment & ToolboX suite (https://qunex.yale.edu/) that enables cloud-based containerized neuroimaging biomarker analytics (42) and iv) DataOrc, an internally developed software tool designed to streamline data management and allow flexible interoperability between systems.

To ensure seamless electronic data capture (EDC), all potential participants were assigned a study number at the time of consent using a Pseudo Global Unique Identifier (PGUID) generated by the NIMH Data Archive (NDA). All eligible participants were also assigned a Simple ID which was provided once the subject was randomized, and was used for the pharmacy label, behavioral data collection, physiological data collection, and eye-tracking data collection.

Figure 3 provides an overview of the data capture system for the TRANSCENDS study. This system ensures consistency and compliance as per protocol and enables seamless integration of all components of the clinical trial (Sponsor and Monitoring Agencies, Pharmacy, Research Team, Site Clinical Staff, Patient, Direct Data Capture, REDCap, XNAT)

**Fig. 3.**
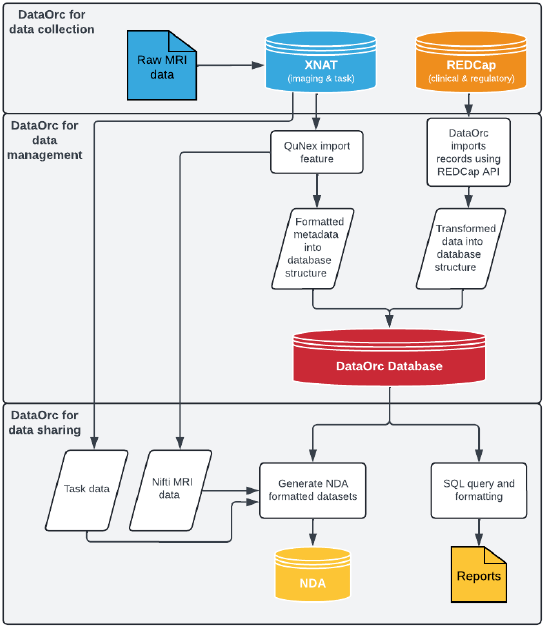
DataOrc Overview: From data collection to data sharing. (Top) DataOrc for data collection: The DataOrc XNAT module is used to upload Raw MRI data into XNAT. (Middle) DataOrc for data managment: A DataOrc database is created and integrates imaging metadata and REDCap assessments (clinical, demographics and regulatory data). Raw MRI DICOMs are converted to NIfTI format for upload to NDA, using QuNex. The metadata from the NIfTI conversion is put into the DataOrc database. For REDCap, the DataOrc REDCap module is used to import records via the REDCap API, transform them into a database structure, and store them in a DataOrc database. (Bottom) DataOrc for data sharing: DataOrc NDA module is then used to generate NDA formatted datasets (CSVs) for upload / sharing to NDA. This module integrates information stored in the DataOrc database including task data and MRI in NIfTI format downloaded from XNAT using the DataOrc XNAT module. DataOrc is then used to generate reports for seamless data sharing with sponsor, study team and external agencies (e.g. DSMB, NIH, FDA).

**Fig. 4.**
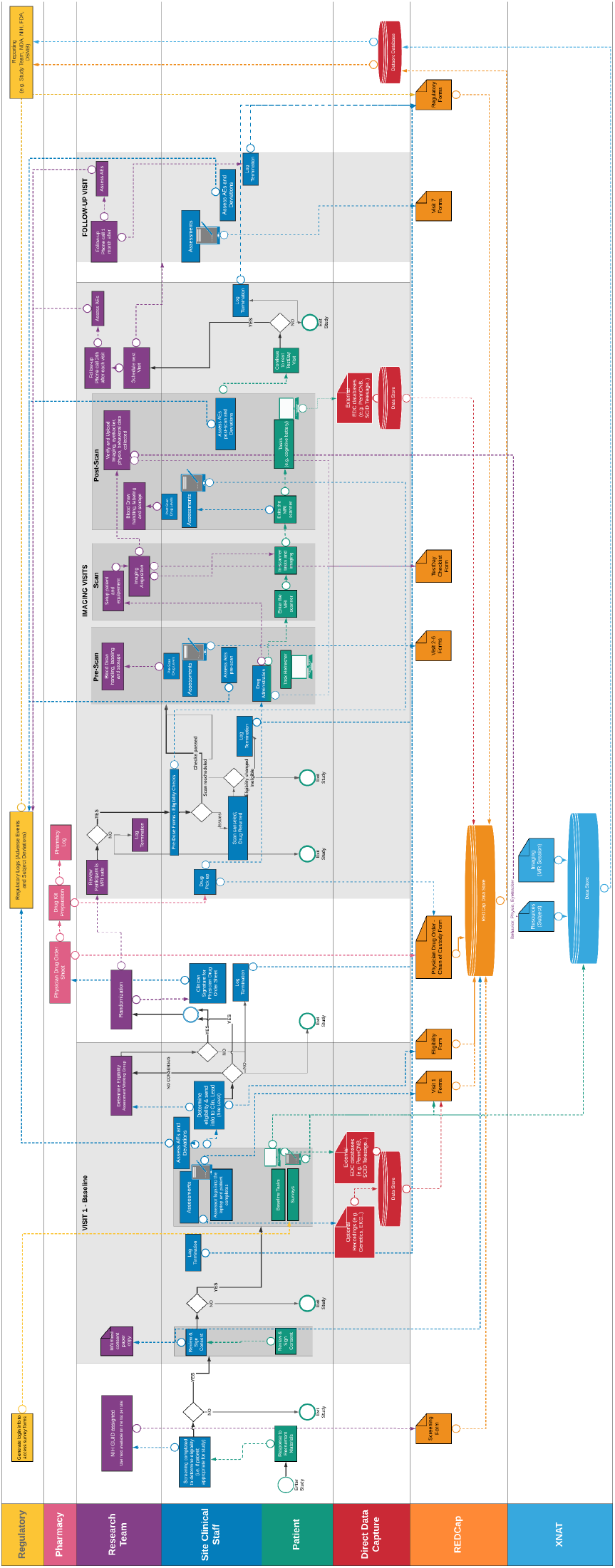
Overview of the data capture system for the TRANSCENDS study. This inter-operable informatics infrastructure enables seamless integration of all components of a clinical trial. This overview diagram showcases the e-source workflow from the entry point of a patient responding to recruitment materials, through all actions done within each study visit, to the closeout of the study with reporting to regulatory agencies (e.g. NIH, NDA, FDA, DSMB). Circles represent entrance/exit of the workflow. Rectangles represent actions (e.g. assessments) done by the component. Dashed arrows represent flow of information within and between clinical trial components (e.g. assessments collected on tablets during Visit 1 are directly captured in the CFR compliant database). Diamonds represent decisions that have to be made by the component (e.g. Eligibility YES or NO decided by the site clinical staff component). Solid arrows represent path of the decision made. Form symbols represent the input of data objects (e.g. specific forms prepared within REDCap). Grey rectangles specify the different visits the patients will go through, e.g. Baseline (Visit 1), Test Days (Visits 2-6) and Follow-up (Visit 7). All actions located in the grey rectangles are done within this visit. Colors for each clinical trial component are Yellow: Regulatory compliance; Pink: Pharmacy; Purple: Research Team; Dark Blue: Site Clinical Staff; Green: Patient facing interactions; Red: External Data Capture Components; Orange: REDCap system; Light Blue: XNAT system.

### Integration of imaging and clinical endpoints into a unified infrastructure

#### Inter-operable and Unified Infrastructure

An in-house software tool, DataOrc, was developed in order to streamline data management and allow flexible transformations and integration between systems. DataOrc facilitates data flow to and from XNAT by integrating with the XNAT REST API. Together, this set-up enables robust support of DICOM uploading and importing, downloading of raw or processed neuroimaging data, and integration of information required for NDA upload. Additionally, DataOrc imports data from the REDCap API and transforms it into a structured meta-data database that can be queried for quality control and automated reporting, and can facilitate exports for upload to the NDA. The database structure definition and transformation steps utilize unified configuration files to define the relationships between fields in each system, allowing for flexible configuration to a study’s specific instrument design and overall data organization. Unlike REDCaps entity-attribute-value model, DataOrc organizes data into structured tables for efficient SQL queries. Distributed as a Rust binary, it ensures easy deployment with minimal dependencies while maintaining data security by querying only necessary fields and preventing PHI release.

### Regulatory considerations for a primary functional neuroimaging-based readout

The inclusion of a drug in the TRANSCENDS study requires compliance with the FDA’s Investigational New Drug (IND) application process. This protocol ensures participant safety and data integrity throughout the clinical trial, adhering to the standards set by the Code of Federal Regulations (CFR). The objectives for the TRANSCENDS data management framework are to:

1. Build an Electronic Data Capture (EDC) system, replacing paper-based data collection;
2. Design the EDC to facilitate remote monitoring;
3. Ensure regulatory compliance of the EDC system with 21 CFR Part 11;
4. Enhance data sharing capabilities;
5. Integrate all features into a unified framework.

A 21 CFR Part 11 compliant REDCap system will serve as the core of this framework.

#### 21 CFR Part 11 and HIPAA compliance

The security principles of **confidentiality, integrity, and availability** are the foundation of compliance frameworks, including 21 CFR Part 11 and HIPAA. Both regulations draw from established security frameworks, such as those provided by the National Institute of Standards and Technology (NIST) (79) and the International Organization for Standardization (ISO) (80), to ensure that systems handling sensitive data adhere to rigorous standards of trustworthiness and security.

21 CFR Part 11 refers to a set of regulations from the Food and Drug Administration (FDA) that govern the use of electronic records and signatures (81). It specifically applies to records created, modified, or stored electronically.

21 CFR Part 11, Section 11.1(a) further states that electronic records in compliance with 21 CFR Part 11 criteria shall be considered by the agency to be “trustworthy, reliable, and generally equivalent to paper records” (82). In order to achieve 21 CFR Part 11 compliance, documentation needs to be created and executed, as well as controls implemented. Documentation and controls are critical pieces of compliance and security because they:

- Provide evidence of compliance during regulatory inspections **(integrity and availability)**;
- Ensure the system is properly installed, tested, and validated **(integrity and availability)**;
- Maintain the integrity, reliability, and security of electronic records and signatures **(confidentiality, integrity, and availability)**;
- Reduce risks associated with system failures, human error, and data breaches **(confidentiality, integrity and availability)**;
- Create a clear audit trail for traceability and accountability **(confidentiality, integrity, and availability)**.

Key documentation includes an Installation Qualification Plan, System Test Plan, Validation Plan, and Compliance Determination document (82). These documents ensure compliance and are reviewed during inspections. Though these documents are not explicitly referred to in the regulation, they are generally considered required for compliance based off of FDA-issued guidance documents and the previously mentioned security frameworks (81). Part 11 requires three levels of control:

- **Administrative**: Policies and use of electronic signatures **(confidentiality, integrity, and availability)**;
- **Procedural**: Standard Operating Procedures (SOPs) for system use **(integrity and availability)**;
- **Technical**: Software functions ensuring record reliability and integrity **(confidentiality and integrity)**.

The Yale Center for Clinical Investigation (YCCI) 21 CFR Part 11 compliant REDCap is a closed system. A system is closed when the system is under the control of persons who are responsible for the electronic records managed by this system (11.3(b)(4)). A closed system is essential for ensuring compliance with 21 CFR Part 11 because it provides the necessary control, security, and reliability to protect electronic records and signatures. See 21 CFR Part 11 regulation (11.10) (82) for the list of requirements of a closed system. The Health Insurance Portability and Accountability Act (HIPAA) consists of three main components:

- **Privacy Rule:** Patients have the right to access their medical records, and to request corrections. Healthcare providers must obtain patient consent before sharing PHI.
- **Security Rule:** Establishes national standards for securing electronic PHI (ePHI). Requires covered entities to implement administrative, physical, and technical safeguards to protect ePHI.
- **Breach Notification Rule:** Requires covered entities to notify affected individuals, the Department of Health and Human Services (HHS), and sometimes the media in the event of a breach of unsecured PHI.

Both REDCap and XNAT are HIPAA compliant, ensuring the confidentiality, integrity, and availability of protected health information.

#### Data Management

##### Electronic Case Report Form (eCRF) Design and Development

- **User Acceptance Testing (UAT)**: REDCap and XNAT have separate UAT plans for 21 CFR Part 11 compliance. Testing was conducted in controlled environments before final implementation.
- **Go-Live Process**: After training, sites (Yale, Penn, SBU, Columbia/RFMH) validated the platforms prior to proceeding to production.
- **eCRF Guidelines**: All details were outlined in the MOPs and SOPs for the study. Trial support materials were stored in a secured Git repository. The eCRF process includes identifying necessary database modifications, revising and testing updates, and implementing approved amendments. Once testing is completed, the data management team updates all relevant materials and releases the changes to the study team.

#### Database Privileges and Access Management

Access to REDCap and XNAT is role-based, adhering to the principle of least privilege. Each site is assigned a specific Data Access Group (DAG) that limits the user to only view the record ID added to a specific DAG, while XNAT restricts access to site-specific projects.

#### *Data Monitoring*. Data Quality and Control Checks

To ensure accurate data collection and transfer, checklists within REDCap guide sites and support timely validation. Open records in REDCap are reviewed weekly, allowing the team to monitor new entries, initiate queries, and assign them to site-specific coordinators. Queries address incomplete, ambiguous, inconsistent, or missing data, such as skipped items or critical forms like termination records. REDCap’s built-in validation rules (e.g., required fields, out-of-range values, and skip patterns) enforce data integrity, and sites areresponsible for resolving queries promptly. Outstanding issues are discussed during Regulatory Working Group (WG) calls.

For enhanced validation, the REDCap database was transformed into a structured SQLite relational database, enabling advanced consistency checks using DataOrc. This allows implementation of complex validation logic, handling protocol changes, and ensuring alignment with NDA data structures. To verify neuroimaging data integrity, a protocol validation framework on XNAT automatically checks uploaded sessions against expected scan templates, flagging discrepancies. The protocol validation framework queries the XNAT API to retrieve detailed information for all the scans in a session, including the series description, the number of frames and files, and some additional fields in the DICOM headers. This is compared to a template with the expected series description and number of files for each scan. Weekly reviews are done to assess neuroimaging, task, and physiological data, with issues logged as REDCap queries. Common concerns include naming inconsistencies and mismatches between uploaded data and REDCap records. Site-specific coordinators address queries, which are then resolved by the data management team once corrected.

Raw image quality is also assessed during this time using the QuNex container to generate reports for each session.

#### Reports

Automatic reports are created in REDCap to extract weekly recruitment numbers, and the status of reporting adverse events (AEs) or deviations. Alerts for new enrollments and drug visits are also set up to allow the sponsor to confirm accurate eligibility and that visits went smoothly. Reports are generated by DataOrc and used to create annual reports (FDA / NIH / Cerevel) and DSMB reports.

### Seamless Data Sharing

#### Intraoperability with NIMH Data Archive (NDA)

NIH-funded studies must upload data to the NDA biannually, necessitating automated, scalable tools for data processing and formatting. We extended DataOrc into a repeatable, reproducible, and fully automated system that generates NDA-formatted datasets directly from source data capture systems. DataOrc uses YAML configuration files to map relationships between REDCap, XNAT, and the NDA data dictionary, handling data encoding transformations and derived calculations. Unlike existing tools, DataOrc offers a flexible, extensible solution for studies using REDCap and XNAT, eliminating manual operations while ensuring automated error checking and verification. For neuroimaging uploads, DataOrc supports both DICOM and NIfTI formats, generating data packets and CSV files with acquisition metadata. It also integrates XNAT-derived information into NIfTI uploads, enriching metadata beyond standard NIfTI headers and JSON sidecar files.

#### Harmonization across sites

As a multi-site neuroimaging initiative, the TRANSCENDS study prioritized validation of neuroimaging data acquisition, transfer, and processing procedures prior to the onset of clinical data collection. To ensure cross-site consistency, members of the research team underwent scanning procedures at each participating site. This within-subject design enabled direct comparisons of imaging data, as well as behavioral and eye-tracking measures, across different scanners and environments. We deliberately chose this approach over the exclusive use of imaging phantoms, as phantoms cannot simulate task-based functional imaging or behavioral data collection. Moreover, we selected trained research personnel to undergo scanning at all sites, rather than relying on early patient data, to allow controlled, within-subject evaluation of acquisition protocols. This strategy provided a reliable means of assessing and adjusting for potential sources of variability prior to participant enrollment.

Given the inclusion of both GE and Siemens MRI scanners in the TRANSCENDS study, harmonization of data acquisition parameters is essential. Differences in scanner manufacturer, model, and software introduce known challenges in multi-site imaging research. To address this, standardized acquisition protocols were developed and are summarized in Table 3. Ongoing scanner performance is also monitored via regular phantom scans to ensure longitudinal stability.

**Table 3.** Harmonization of imaging sequences.

|  | Matrix | Slices | FOV | % FOV (phase) | Resolution (mm) | TR (ms) | TE (ms) | TI (ms) | Flip Angle (deg) | Echo spacing (ms) | Bandwidth (Hz/Px) | Parallel Imaging | Multiband Acceleration | Phase Partial Fourier | Diffusion Directions | b-values |
| --- | --- | --- | --- | --- | --- | --- | --- | --- | --- | --- | --- | --- | --- | --- | --- | --- |
| T1w | 320 x 300 | 208 | 300 x 320 | 93.75 | 0.8 x 0.8 x 0.8 | 2400 | 2.22 | 1000 | 8 | 7.5 | 220 | 2x | Off | Off | N/A | N/A |
| T2w | 320 x 300 | 208 | 300 x 320 | 93.75 | 0.8 x 0.8 x 0.8 | 3200 | 563 | N/A | 120 | 3.52 | 745 | 2x | Off | Allowed | N/A | N/A |
| SpinEchoFieldMap | 104 x 104 | 3 | 936 x 936 | 100 | 2.0 x 2.0 x 2.0 | 8000 | 66 | N/A | 90 | 0.58 | 2290 | Off | Off | Off | N/A | N/A |
| fMRI TASK | 104 x 104 | 770 | 936 x 936 | 100 | 2.0 x 2.0 x 2.0 | 800 | 37 | N/A | 52 | 0.58 | 2290 | Off | 8 | Off | N/A | N/A |
| fMRI REST | 104 x 104 | 770 | 936 x 936 | 100 | 2.0 x 2.0 x 2.0 | 800 | 37 | N/A | 52 | 0.58 | 2290 | Off | 8 | Off | N/A | N/A |
| fMRI CHECKERBOARD | 104 x 104 | 400 | 936 x 936 | 100 | 2.0 x 2.0 x 2.0 | 800 | 37 | N/A | 52 | 0.58 | 2290 | Off | 8 | Off | N/A | N/A |
| ASL | 64 x 63 | 8 | 896 x 896 | 100 | 1.5 x 1.5 x 3.0 | 4600 | 17.92 | N/A | 180 | 0.5 | 2695 | Off | Off | Off | N/A | N/A |
| Diffusion 1 | 140 x 140 | 99 | 1400 x 1400 | 100 | 1.5 x 1.5 x 1.5 | 3230 | 89.2 | N/A | 78 | 0.69 | 1700 | Off | 4 | 6/8 | 98 | 3000 |
| Diffusion 2 | 140 x 140 | 100 | 1400 x 1400 | 100 | 1.5 x 1.5 x 1.5 | 3230 | 89.2 | N/A | 78 | 0.69 | 1700 | Off | 4 | 6/8 | 99 | 3000 |

|  | Matrix | Slices | FOV | % FOV (phase) | Resolution (mm) | TR (ms) | TE (ms) | TI (ms) | Flip Angle (deg) | Echo spacing (ms) | Bandwidth (Hz/Px) | Parallel Imaging | Multiband Acceleration | Phase Partial Fourier | Diffusion Directions | b-values |
| --- | --- | --- | --- | --- | --- | --- | --- | --- | --- | --- | --- | --- | --- | --- | --- | --- |
| T1w | 320 x 320 | 224 | 320 x 320 | 90 | 0.8 x 0.8 x 0.8 | 2373.09 | 3.472 | 1000 | 8 | 0 | 195.312 | 2x | Off | Off | N/A | N/A |
| T2w | 320 x 320 | 224 | 320 x 320 | 90 | 0.8 x 0.8 x 0.8 | 3202 | 59.931 | N/A | 90 | 0 | 390.625 | Off | Off | Off | N/A | N/A |
| SpinEchoFieldMap | 108 x 108 | 216 | 108 x 108 | 100 | 2.0 x 2.0 x 2.0 | 8000 | 60 | N/A | 90 | 0.624 | 4629.63 | Off | Off | Allowed | N/A | N/A |
| fMRI TASK | 108 x 108 | 55440 | 108 x 108 | 100 | 2.0 x 2.0 x 2.0 | 800 | 34 | N/A | 52 | 0.64 | 4629.63 | Off | 8 | Allowed | N/A | N/A |
| fMRI REST | 108 x 108 | 55440 | 108 x 108 | 100 | 2.0 x 2.0 x 2.0 | 800 | 34 | N/A | 52 | 0.64 | 4629.63 | Off | 8 | Allowed | N/A | N/A |
| fMRI CHECKERBOARD | 108 x 108 | 29664 | 108 x 108 | 100 | 2.0 x 2.0 x 2.0 | 800 | 34 | N/A | 52 | 0.64 | 4629.63 | Off | 8 | Allowed | N/A | N/A |
| ASL | 512 x 8 | 64 | 128 x 128 | 100 | 1.875 x 1.875 x 4.0 | 4805 | 52.8 | N/A | 111 | 0 | 976.562 | 2x | Off | Off | N/A | N/A |
| Diffusion 1 | 140 x 140 | 8019 | 140 x 140 | 100 | 1.7143 x 1.7143 x 1.7 | 4900 | 78.4 | N/A | 90 | 0.704 | 3571.43 | Off | 3 | Allowed | 98 | 3000 |
| Diffusion 2 | 140 x 140 | 8100 | 140 x 140 | 100 | 1.7143 x 1.7143 x 1.7 | 4900 | 78.4 | N/A | 90 | 0.704 | 3571.43 | Off | 3 | Allowed | 99 | 3000 |

This rigorous approach to cross-site harmonization enhances the integrity of the imaging and behavioral data, supporting robust analyses across diverse clinical and scanner environments.

#### Cloud-based containerized neuroimaging biomarker analytics

All neuroimaging data for this study will be processed in the QuNex neuroimaging suite (42). QuNex is an integrated neuroimaging environment explicitly architected for scale and high throughput of big data, and stress-tested via processing of >10,000 multi-modal imaging sessions across scanner vendors (42). The QuNex Container features leading community tools, including dcm2niix (83), FSL (84), Connectome Workbench (85), Human Connectome Project (HCP) Pipelines (86), PALM (87), and FreeSurfer (88, 89), along with Octave (90) and R Statistical Environment (91) all packaged within the container. For a full list of featured software, see QuNex documentation. QuNex is optimized for cloud-based deployment and enabled for seamless XNAT interoperability via the XNAT container plugin. A turnkey engine enables frictionless deployment of entire pipelines within high-performance compute environments (e.g. SLURM) through a flexible scheduler system via a single command line call. We will deploy HCP-style pipelines for the processing and analysis of anatomical, BOLD functional connectivity, and structural connectivity data in a study-specific XNAT project. Specifically, the XNAT architecture supports the integration and databasing of all MR data uploaded directly from each of the sites with full HIPAA-compliance .

### Analytic Strategy for biomarker readout

TRANSCENDS aims to identify neural biomarkers and mechanisms underlying cognitive deficits in schizophrenia to guide targeted treatments. The statistical analysis plan, approved by the biomarker and neuroinformatics working groups and the study statistician, ensures that the methods are suitable for extracting meaningful biomarker data and rigorous, reproducible results. For the full analytics plan, refer to https://clinicaltrials.gov/study/NCT04457310.

One primary outcome and six secondary outcomes will be assessed:

- Primary Outcome: Spatial working memory neural circuit BOLD signal change in response to CVL-562. A general linear model will estimate hemodynamic response functions (HRFs) across 20 timepoints for each task condition. A 3-way interaction (dose *×* time-point *×* task condition) will be tested in spatial working memory regions using an independent mask from prior work (44). Significant voxels will be identified using non-parametric statistics and voxel-wise corrections for multiple comparisons.
- Secondary Outcomes:
  - Identification of the proportion of participants with a BOLD signal response to CVL-562. A binary outcome will be calculated for each individual based on the dose x timepoint x task condition analysis of the working memory BOLD signal.
  - Spatial working memory performance change. Performance will be measured as the angular difference between the target presentation and the participants response, and analyzed using repeated measures ANOVA (dose x timepoint x task condition).
  - Change in BOLD signal regression with trial-by-trial spatial working memory performance. A general linear model will assess the association between sWM performance and task-evoked BOLD signal, incorporating trial-by-trial sWM performance as a covariate. A dose Œ task condition interaction with the sWM precision covariate will indicate drug-related BOLD signal changes associated with sWM performance.
  - Task-based functional connectivity changes between the fronto-parietal control network (FPCN) and the rest of the brain. Connectivity will be assessed using FPCN as a seed region, analyzing dose-dependent effects on covariation with all other greyordinates during the sWM delay period. Connectivity metrics will undergo second-level random effects analyses to evaluate FPCN functional integrity across task conditions and drug doses.
  - Global brain resting-state connectivity (GBC) changes. Dose-dependent effects of CVL-562 on GBC will be assessed using the data-driven GBC metric, which evaluates connectivity from each voxel to all others. Individual GBC will be entered into second-level random effects analyses.
  - Correlations between transcriptomic maps and CVL-562-induced GBC changes. Resting-state GBC maps will be correlated with gene expression data related to psychotic diseases from the Allen Human Brain Atlas to determine if GBC changes align with regions enriched for schizophrenia-related gene expression (92).

An exploratory endpoint will be examined to test whether genetic variants are related to BOLD signal changes in response to CVL-562.

## Discussion

The Translational Neuroscience & Computational Evaluation of a D1R Partial Agonist for Schizophrenia (TRANSCENDS) study is intended to accelerate the development of a therapeutic agent by establishing its dose-related pharmacodynamic effects on a neuroimaging biomarker. We implemented a cognitive paradigm explicitly designed to capture a translational micro-circuit mechanism underlying spatial working memory in schizophrenia patients. This allows us to test if D1R/D5R partial agonist (CVL-562) induces a dose-dependent restoration of neural tuning measured via fMRI in humans. This biomarker was selected on the basis of a translational and computational understanding of prefrontal micro-circuitry and a mechanistic understanding of the role of D1R/D5Rs in schizophrenia. To support this brain-based clinical trial, we developed an automated and scalable framework that combines a rigorous regulatory framework with automated quality assurance, interoperable electronic data capture, validated systems integration, seamless data sharing, harmonization across sites and analytics for biomarker readout. We hope that TRANSCENDS, with its brain-based outcome measure and rigorous regulatory framework, could provide a model for future neuroimaging studies aimed at identifying novel treatment mechanisms in human participants.

## Future plans

We have begun to expand this infrastructure to other multi-site neuroimaging studies, including those from the Accelerating Medicines Partnership Schizophrenia (AMP SCZ) (93). Using this framework in the large-scale Psychosis Risk Outcomes Network (ProNET, PI:Woods) will further grow the ability of this infrastructure to handle different types of data (EEG, actigraphy, etc) and adhere to international data regulatory standards (GDPR). In the related study, Psychosis Risk Outcomes Compound Assessment Network (ProCAN), measuring the effects of a study drug on brain-based biomarkers will be central to the study design, and the use of our framework in ProCAN aligns well with the goal of TRANSCENDS to develop therapeutics using mechanistically-guided outcomes. Both studies emphasize the need for modular, scalable frameworks that adhere to rigorous regulatory standards, such as 21 CFR Part 11, HIPAA and/or GDPR compliance, to facilitate secure and interoperable data sharing across sites. We hope this optimized ecosystem eases the regulatory burden of future mechanistically-guided clinical trials, thereby accelerating treatment development in psychiatry and beyond.

## Data Availability

This protocol paper does not include primary data; data will be made available upon completion of the study.

## Acknowledgments

Cerevel provided the drug CVL-562 for this trial.

## TRANSCENDS Group members

***Yale University School of Medicine.*** Philisha Abrahim, Ryan Aker, Christopher Artabane, Anahita Bassir Nia, Samuel Brege, Laura Cadavid, Terry Camarro, Jessica Campbell, Deepak D’Souza, Alyssa Gateman, Theresa Goddard, Sandra Gomez-Luna, Mackenzie Griffin, Ralitza Gueorguieva, Aarti Gupta, Brittany Harris, Mara Heneks, Julie Holub, Tesheia Johnson, Kangjoo Lee, Alicia Moroyoqui, John Murray, Prashant Patel, Jasmine Patel, Rajiv Radhakrishnan, Ali Rashid, Helen Seow, Vinod Srihari, Sharath Vallabhajosyula, Kimberly Vasquez

***Columbia University, College of Physicians and Surgeons / New York State Psychiatric Institute.*** James Gangwisch, Jayda Melnitsky, Renu Nandakumar, Joanne Yoon

***Stony Brook University Renaissance School of Medicine.*** Sameera Abeykoon, Topaz Baumvoll, Kelly Bobchin, Richard Connor, Jean-Paul Farhat, Francesca Giammanco, Thomas Jaworski, Jisoo Kim, Ryan O’Connor, Tina Yip

***University of Pennsylvania Perelman School of Medicine***. Mark Elliott, Nina Laney, Kosha Ruparel, Sage Rush, Lyndsay Schmidt

## Contributors

Conceptualization: A.A., J.H.K., R.E.G., A.A.D., and J.A.L. Funding acquisition: A.A. and J.H.K. Project administration: C.F., Y.T.C., N.S., and Z.T. Writing original draft: C.F., Z.T., and Y.T.C. All authors reviewed and approved the final manuscript.

